# WearGait-PD: An Open-Access Wearables Dataset for Gait in Parkinson’s Disease and Age-Matched Controls

**DOI:** 10.1101/2024.09.11.24313476

**Authors:** Anthony J. Anderson, David Eguren, Michael A. Gonzalez, Naima Khan, Sophia Watkinson, Michael Caiola, Siegfried S Hirczy, Cyrus P. Zabetian, Kelly Mills, Emile Moukheiber, Laureano Moro-Velazquez, Najim Dehak, Chelsie Motely, Brittney C. Muir, Ankur Butala, Kimberly Kontson

**Author notes:** corresponding author: Kimberly Kontson. These authors contributed equally to the manuscript (Anthony J. Anderson, David Eguren, Michael A. Gonzalez).

## Abstract

Wearable movement sensors are powerful tools for objectively characterizing and quantifying movement. They enhance the precise characterization of gait, balance, and motor symptoms in Parkinson’s disease and related disorders, facilitating in-clinic and remote assessments, disease management, and therapeutic intervention development. Access to high-quality data from these sensors can accelerate discoveries in this clinical population. The WearGait-PD open-access dataset contains raw inertial measurement unit (IMU) and sensorized insole data from individuals with PD and age-matched controls, synchronized to a gait walkway reference system. IMU data include 3-degree of freedom (DOF) acceleration, rotational velocity, magnetic field strength, and orientation for each of 13 sensors on the participant’s body. Sensor insole data include absolute pressure from 16 sensors in each insole and 3-DOF acceleration and rotational velocity. Walkway data include 2D position and relative pressure for each active sensor during every footfall. Frame-by-frame annotation of participant actions during gait and balance tasks was incorporated using synchronized video cameras. All data were associated with demographic information and clinical evaluations (e.g., medications, DBS-status, MDS-UPDRS scores).

## Background & Summary

Parkinson’s disease (PD) is a progressive neurodegenerative disorder that often impairs gait and balance, resulting in reduced mobility and independence, and diminished quality of life^1-3^. Wearable movement sensors have emerged as powerful tools for objectively characterizing and quantifying movement. These offer the potential to enhance the precise characterization of gait, balance, and motor symptoms in Parkinson’s disease (PD) and related disorders, facilitating in-clinic and remote assessments, disease management, and therapeutic intervention development^4-7^. In contrast, current clinical assessments of gait and balance are semi-quantitative but remain largely qualitative, relying on subjective observations and patient reports^8-10^. Wearable sensors promise to improve care by enabling quantitative assessments as part of routine clinical practice^11^, remote patient monitoring^12,13^, and objective endpoint measures for the regulatory evaluation of new drugs and medical devices^14^. By providing continuous, high-resolution data on mobility-related health, these sensors and their associated algorithms can identify motor signs of PD, track disease progression, and offer a more complete picture of an individual’s functional status^15-17^. This data-driven approach can potentially transform PD treatment, ultimately improving patient outcomes and quality of life.

Despite the potential of wearable movement sensors to improve the treatment of PD, their translation and widespread clinical adoption have been limited due to various scientific, technical, economic, and regulatory issues^18-20^. However, efforts are underway across academia, the medical device industry, and regulatory bodies to overcome these limitations. For instance, academic-led consortiums, such as Mobilise-D, are working to establish, verify, and validate digital mobility outcomes, creating a framework for their use in clinical settings^21^. Medical device developers have created wrist-worn devices that monitor the presence of upper limb symptoms of PD and incorporate these measurements into clinician-facing dashboards in an aim to improve treatment decisions^22,23^. Additionally, the FDA has issued guidance on using digital health technologies for remote data acquisition in clinical investigations^24^ and participates in the Digital Health Measurement Collaborative Community (DATAcc) by the Digital Medicine Society, while the European Medicines Agency recently approved a digitally-derived measure for use in Duchenne Muscular Dystrophy clinical trials^14^. These activities indicate a growing recognition by regulatory organizations of the transformative potential of the technology.

The need for high-quality, diverse, and well-annotated data remains a significant obstacle to the development, validation, and clinical translation of wearable sensor digital health technologies. As digital health technologies rely on a combination of computing platforms, sensors, and algorithms^24^, data are a fundamental requirement at every stage of development, from prototyping through clinical trials^25^. However, obtaining high-quality movement data from people with PD is challenging due to the high costs associated with data collection, the fragility of the population, and the necessity for skilled clinicians to score and annotate the data. These barriers create a development landscape where only well-funded organizations, public or private, have the resources to participate in the innovation process, limiting the diversity of ideas and approaches explored.

Open-access datasets offer one solution to the challenge posed by the lack of data in the development of wearable technologies for PD. By providing high-value clinical datasets to the research and development communities, open-access data efforts can democratize the innovation process, enabling a wider range of researchers and developers to contribute to advancing these technologies. Open-access datasets also promote reproducibility by allowing researchers and developers to validate their findings independently and with diverse data. While several open-access wearable sensor and smartphone datasets for PD already exist^26-33^, their overall utility may be limited by: (1) a primary focus on wrist-based monitoring, which may not capture the full spectrum of gait and balance impairments, (2) a lack of associated clinical information, such as disease severity scales and medication status, (3) insufficient contextual information surrounding the data, such as time-series descriptions of specific locomotor activities, and (4) a lack of reference measurements for the analytical validation of algorithms^25^. Addressing these limitations is crucial for streamlining the integration of wearable sensors for clinical use in PD.

In response to these limitations, we present a large, multi-site, multi-modal open dataset (WearGait-PD) of older adults with and without PD, engaged in both structured and semi-structured gait and balance tasks while wearing a full-body suite of sensors (Figure 1). This dataset fills several vital gaps in the field, as it: (1) provides data from a full-body sensor suite of inertial measurement units and sensorized insoles, (2) includes comprehensive clinical information, such as Movement Disorder Society – Unified Parkinson’s Disease Rating Scale (MDS-UPDRS) scores and demographic details for each participant, (3) offers frame-by-frame, human-readable annotations applied by two expert reviewers based on video assessment, with clinician annotation of PD-specific symptoms such as freezing of gait, and (4) incorporates ground truth measurements from a pressure-sensing walkway for independent validation of digitally-derived metrics. By making this dataset publicly available to the research and medical product development communities, we aim to accelerate innovation and ultimately access to effective digital health technologies for assessing gait and balance in PD.

**Figure 1:**
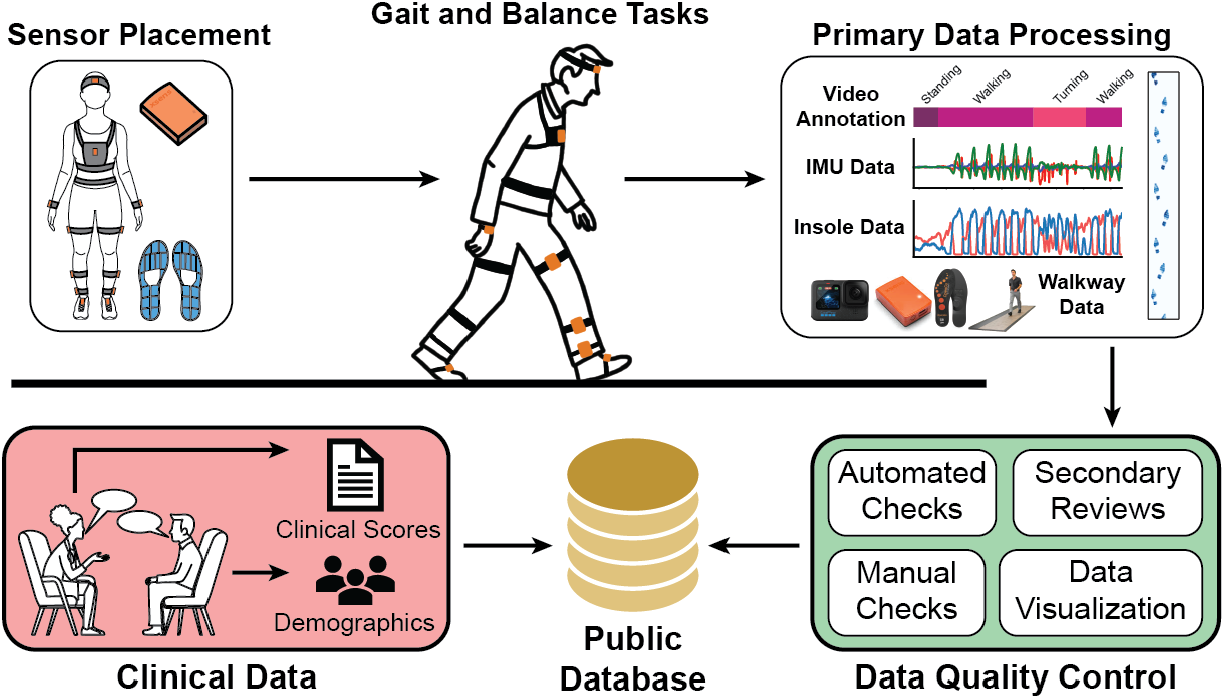
Overview of data workflow for the WearGait-PD dataset. Sensor data were collected via a set of wearable sensors (IMUs and sensorized insoles) and reference systems (cameras and a sensorized walkway). The sensor data were processed, time synchronized, visualized, and reviewed before uploaded to the public repository. Clinical data included demographic information and results from clinical tests when possible.

## Methods

### Participants

At the time of submission, 126 participants were included in the dataset. Sixty-one PD participants (25F/36M) with an average age of 66 ± 9 years and modified Hoehn & Yahr score of 2.15 ± 0.48 were included. The total MDS-UPDRS Part III score for these participants ranged from 6-46, with a mean and standard deviation of 24 ± 10. The protocol did not require participants to perform tasks in a specific medication state (i.e., ON or OFF). Instead, the time elapsed since their last PD medication dose was recorded as a proxy. A total of 41 freezing of gait events were captured across five unique PD participants. Within the PD group, 158 stagger events were captured. Sixty-five control participants (40F/25M) with an average age of 76 ± 9 years were also included. Within this control group, 163 staggering events were captured.

### Inclusion/Exclusion Criteria

The study included adults (≥18 years) with physician-diagnosed PD and controls without PD. Exclusion criteria encompassed conditions that could prevent safe task performance, including non-neurological disorders significantly affecting speech, vision, or balance, cognitive or psychological impairments, or pregnancy. These criteria allowed for the inclusion of participants with a wide variety of typical and mildly pathological movement characteristics within the control group (e.g., individuals with essential tremor were not excluded, as we aimed to avoid spectrum bias by having only controls).

### Consent and Ethics

Written informed consent was obtained from all study participants, including permission to publish de-identified data openly. These data were collected under the Johns Hopkins School of Medicine (JHM) IRB00234370 with an IRB Authorization Agreement (95-CDRH-2021-12-18) and VA IRB01702255 with an IRB Authorization Agreement (2023-CDRH-095), for which FDA is the relying institution.

### Sensor Systems

The following sensor systems were used during the collection of data for this study. All study sites used the same acquisition software versions to maintain consistency.

*Movella/Xsens MTw Awinda Inertial Measurement Units (IMUs):* MTw Awinda is a wireless human motion tracking sensor suitable for real-time applications. Each sensor was 47 × 30 × 13 mm. Each sensor communicated wirelessly with one of two base stations. Each base station was connected to a computer running the MT Manager 2019.1.1 acquisition software.

*Moticon OpenGo Sensor Insoles:* Moticon OpenGo is a wireless in-shoe system with Bluetooth™- and Wi-Fi-enabled electronics. Each sensor insole is designed to be used inside a shoe and measures plantar pressure distribution at the sole of foot with sixteen distributed pressure sensors. An on-board sensor at the center of the insole measures acceleration and rotation of the foot along three axes. The insoles slip into the shoes of the participant and a mobile Android application (OpenGo Mobile App Version 03.11.00) was used to control data acquisition.

*ProtoKinetics Pressure Walkway:* The 16-foot x 2-foot ProtoKinetics Zeno™ Walkway Gait Analysis System detects pressure data and uses that data to define foot contacts. The walkway was equipped with over 18,000 individual sensors that each registered 16 different activation levels. Each sensor was 0.5” x 0.5” (1.27 cm x 1.27 cm). The ProtoKinetics Movement Analysis Software (PKMAS) 6.00c3 was used for data acquisition and raw data export.

*GoPro HERO10 Black Video Camera:* Two GoPro cameras were used in this study to film the participants from the frontal and sagittal planes as they completed each task. Each camera recorded video at 50 fps with a resolution of 1080p using either the linear or wide camera lens. The video compression was set to H.264 + HEVC. The cameras were triggered wirelessly via remote control by experimenters during gait and balance tasks.

### Experimental Protocol

#### Study Sites

Data were collected across four study sites: FDA Office of Science and Engineering Labs (FDA), Johns Hopkins Outpatient Center (JHOC), Johns Hopkins Bayview Campus (Bayview), and the VA Puget Sound Health Care System Center for Limb Loss and Mobility (VA-Seattle). Subject identifiers specific to each study site were used to denote the location at which data were collected. The prefix ‘FHC’ is associated with the FDA site, ‘NLS’ is associated with the JHOC site, prefix ‘HC’ is associated with the Bayview site, and prefixes ‘WPD’ and ‘WHC’ are associated with the VA-Seattle site. Staff at each study site were trained on the same equipment and the same experimental protocol was used across all sites.

### Participant Preparation

IMU sensors were attached to the participant’s body at the following locations (see Figure 2):

**Figure 2:**
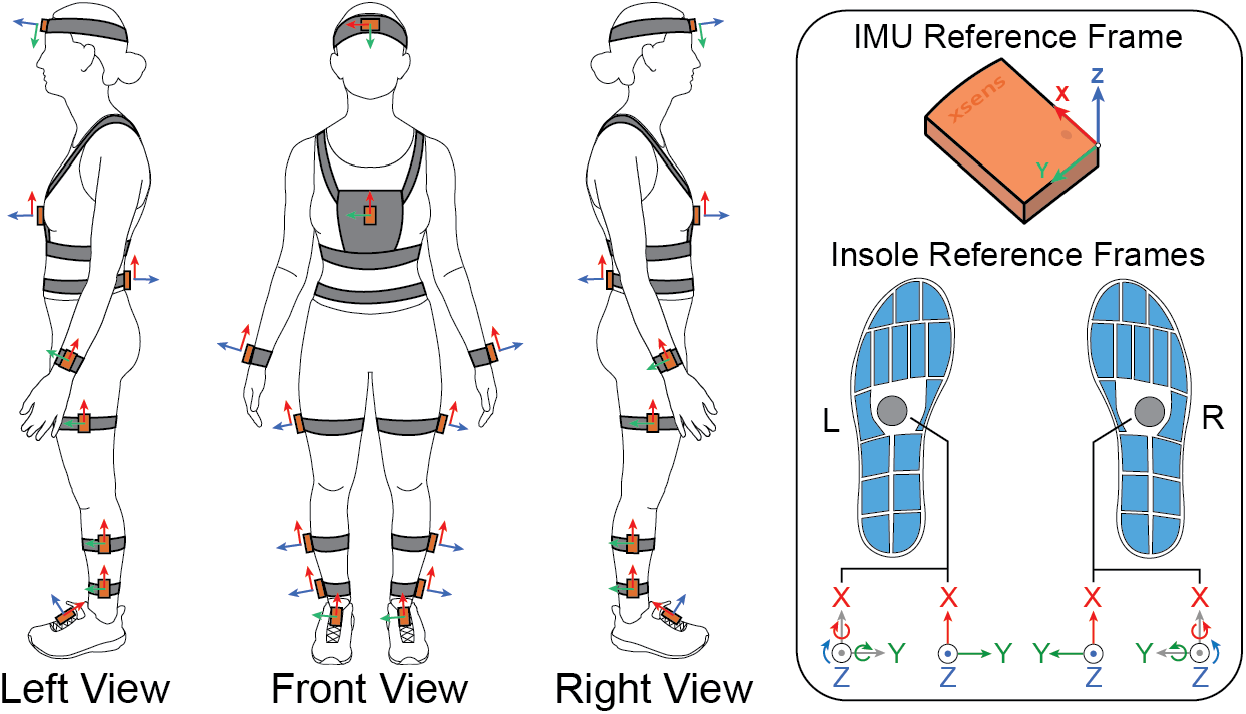
(Left) Locations and orientations of IMU sensors on the body and (Right) the reference frames for both the IMUs and insoles.

- Mid forehead
- Xiphoid process of sternum
- L4/L5 of lower back
- Right/left wrist halfway between the ulnar and radial styloid processes
- Right/left lateral thigh, midway between greater trochanter of femur and head of fibula
- Right/left lateral shank, midway between head of fibula and center of lateral malleolus
- Right/left ankle, just above the lateral malleolus
- Right/left dorsum of feet 50% of distance between 1st and 5th metatarsal bases and 50% of distance between distal end of 3rd metatarsal and center of anterior aspect of talus.

The sensor locations and orientations were standardized and consistently applied across all participants and study sites. Each participant was fitted with Moticon OpenGo sensor insoles placed directly in the shoes.

### Tasks

Once prepped with all study equipment, participants were video-recorded and asked to perform specific tasks. Most tasks required the participants to make several passes along the 16-foot pressure walkway, where a pass constituted walking one length of the walkway. A list of the tasks performed is below, with the abbreviations used in data file names in parentheses. Throughout these task descriptions, the term ‘mat’ is synonymous with ‘walkway’.

- SelfPace (SP): Participants started approximately 5 feet off the mat, walked at a self-selected pace across the mat and approximately 5 feet off the other side of the mat. Participants turned around off the mat and repeated this pass until a total of 4 passes were made.
- HurriedPace (HP): Participants started approximately 5 feet off the mat, walked at a hurried pace across the mat and approximately 5 feet off the other side of the mat. Participants turned around off the mat and repeated this pass until a total of 4 passes were made.
- SelfPace_mat (SPm): Participants repeated the SelfPace walking task as above but remained on the mat the entire time. Participants started on the mat, walked at a self-selected pace across the mat, stopped, and turned around at the other end of the mat. Participants repeated this until a total of 4 passes were made.
- HurriedPace_mat (HPm): Participants repeated the HurriedPace walking task as above but remained on the mat the entire time. Participants started on the mat, walked at a hurried pace across the mat, stopped, and turned around at the other end of the mat. Participants repeated this until a total of 4 passes were made.
- SelfPace_matTURN (SPmT): Participants repeated the SelfPace_mat walking task as above but *were instructed to alternate between left and right turns*. Participants started on the mat, walked at a self-selected pace across the mat, stopped, and turned around at the other end of the mat. Participants repeated this until a total of 5 passes were made so that two turns in each direction were captured.
- TandemGait (TG): Participants made 2 full passes on the mat while doing a tandem gait walk (walking heel to toe). Participants remained on the mat the entire time and turned around naturally after the first pass.
- Timed Up and Go (TUG): Participants started seated in a chair with armrests with their back against the chair. The participant rose from the chair, walked to the other end of the mat at a comfortable, self-selected pace, and turned at a line taped on the mat 3m (9ft 10in) from the chair front. From there, they walked back to the chair, and sat down. Participants were instructed to only use the armrests as needed. Participants did NOT need to cross their arms when standing and sitting. The *front* legs of the chair were placed on the starting line on the mat and turns were made on the mat. The participant completed 3 trials of the TUG test within one continuous recording.
- Balance (B): Participants started this task off the mat, typically near the midpoint of the mat on the side. For each of 6 subtasks, the participant was asked to step onto the mat, complete the balance subtask for 10 seconds, and then step off the mat.
  ∘ Standing with eyes open and feet shoulder width apart
  ∘ Standing with eyes closed and feet shoulder width apart
  ∘ Standing with eyes open and feet together
  ∘ Standing with eyes closed and feet together
  ∘ Standing with eyes open and right foot heel touching the left foot toe
  ∘ Standing with eyes open and left foot heel touching the right foot toe
- SelfPace_doorpat (SPdoorpat): A mock door frame approximately 3 ft x 8 ft was placed across the center of the mat over a pattern of lines perpendicular to the direction of travel. Participants started approximately 5 feet off the mat, walked at a self-selected pace across the mat through the mock door frame and approximately 5 feet off the other side of the mat. Participants turned around off the mat and repeated this pass until a total of 4 passes were made.
- FreeWalk (FW): Participants started approximately 5 feet off the mat, walked at a self-selected pace across the mat and out of the session room into the hallway. Participants proceeded to walk through a defined path within the research facility as they navigated hallways, a small staircase (if available), and sitting down/rising from a chair before returning to the session room. Each study site had a unique free walk path, which is described in greater detail in the Synapse repository.Tasks were completed sequentially in the order listed above.

### Clinical Information and Assessments

In coordination with the clinical collaborator at each study site, information about medication usage and clinical evaluations was obtained from the most recent patient visit when available. If medical records were not available, medication usage and any other relevant medical history were obtained by participant self-report. The MDS-UPDRS was used as the main clinician/patient reported outcome measure. When possible, the MDS-UPDRS evaluation was completed by a trained specialist the day of the session to ensure the evaluation corresponded with the data collected. For instances where the MDS-UPDRS was not captured the day of data collection, the most recent MDS-UPDRS evaluation was used with the date captured denoted in the clinical spreadsheet.

Additional clinical information collected included:

- Time since first diagnosis
- Current medication type and dose
- Time since last medication dose
- Current physical/occupational therapy status and frequency of visits
- DBS (if patient has it); Bilateral vs unilateral; location of electrodes; date of surgery
- Basic demographics (age, gender, sex, height, weight, race)
- Modified Hoehn and Yahr scale score

### Data Processing

*Movella/Xsens MTw Awinda Inertial Measurement Units (IMUs):* During data collection, each MTw IMU internally sampled data at 1000 Hz, applied factory calibration, and low pass filtered at 184 Hz. An onboard stap-down integration algorithm processed these samples, computing orientation and velocity increments that were transmitted wirelessly at 100 Hz. The MT Manager software was used for post-processing, applying the manufacturer’s XKF3-hm Kalman filter to generate 100 Hz outputs of acceleration, angular velocity, orientation, and gravity-compensated (free) acceleration. For a detailed description of the system’s signal processing architecture, refer to the MTw Awinda whitepaper^34^.

*ProtoKinetics Pressure Walkway:* After data collection, the ProtoKinetics software automatically labels groups of active sensor cells as belonging to either the left or right foot. Additional manual preprocessing of the walkway data was completed to ensure left and right footfalls were correctly identified. Partial footfalls on the walkway were labeled as ‘Other’ rather than left or right. After the footfall labeling was completed, a research staff member conducted a review of the labeling to ensure no mistakes or mislabeled footfalls exist.

*Moticon OpenGo Sensor Insoles:* Sensor insole waveforms were reviewed visually. Although infrequent, instances of desynchronization occurred during wireless communication between the sensor insoles and the Moticon acquisition App. This resulted in asynchronous data capture or prolonged recording from one insole even after the ‘Stop’ command was issued. When such desynchronization was detected, the data from the insoles were adjusted—shifted and/or trimmed—within the OpenGo Software to ensure proper alignment between both insoles and to accurately synchronize the cessation of recording with the conclusion of the data stream.

*GoPro HERO10 Black Video Camera:* Manual annotation of the participant’s activity using the video data was also completed. After the data collection session ended, the videos recorded for each trial from the side and front views were combined and deidentified. The result was a single video with both views shown side-by-side, where each view had the participant’s face cropped out. On some occasions, only a single video camera view was available. The primary annotation process of the session videos used the Video Labeler application in MATLAB to provide a frame-by-frame annotation of general and clinical events for the duration of each task video, with clinical events being labeled concurrently with general events.

After the primary annotations were completed, a research staff member conducted a review of the existing annotations to ensure no mistakes or mislabeled events existed. If any clinical events were identified by the research staff during primary annotation or review, those events were reviewed by at least one neurologist collaborator at Johns Hopkins or the VA to confirm or reject the annotation of the clinical event.

Detailed definitions of the general and clinical gait events used in this dataset can be found in the Data [Description – Video Annotation](https://www.synapse.org/Synapse:syn52540892/wiki/625988) section of the Wiki.. These definitions were provided to ensure consistency across annotators.

### Data Synchronization and Alignment

Each task included four main data sources: IMUs, sensor insoles, walkway, and video cameras. The walkway served as the primary clock for all system synchronization and alignment. Upon starting a recording in the PKMAS walkway software, a 3.3 V TTL square wave signal was sent from the ProtoKinetics walkway interface box to the Sync In port of each MTw Awinda base station. The MT Manager software for the MTw Awinda IMUs was configured to start recording upon receipt of the TTL signal from the walkway. Synchronization between the sensor insoles and walkway was achieved by setting the Moticon OpenGo App to produce an audible beep during recording start/stop. The audio signal from the phone was converted to a 3.3 V square wave via custom electronics from Moticon, which was read into the ProtoKinetics walkway interface box and subsequently the walkway acquisition software. Walkway, sensor insole, and IMU systems were all set to record data at 100 Hz.

Two video cameras set up perpendicular and parallel to the direction of participant travel collected data to facilitate frame-by-frame annotation of participant movement. A green light connected to the ProtoKinetics walkway interface box was programmed to turned on for five seconds when recording was initiated. The video cameras started recording prior to the walkway to ensure each video camera captured the green light turning on. The green light was then used to align the two video streams to each other as well as the walkway. For each task, the general order of operations for collecting synchronized data were as follows:

1. Prime IMU acquisition software on computers to accept a sync signal.
2. Start the video cameras using a remote control.
3. Start the trial in walkway acquisition software (which simultaneously starts recording IMU data and triggers green light).
4. Start recording sensor insole data in phone app.

After a participant completed a task, the opposite order of operations was taken to stop recording data:

1. Stop recording sensor insole data in phone app.
2. Stop recording the trial walkway acquisition software (which simultaneously stops the IMU recording).
3. Stop the video cameras using the remote control.

Figure 3 shows a schematic of the overall hardware synchronization approach.

**Figure 3:**
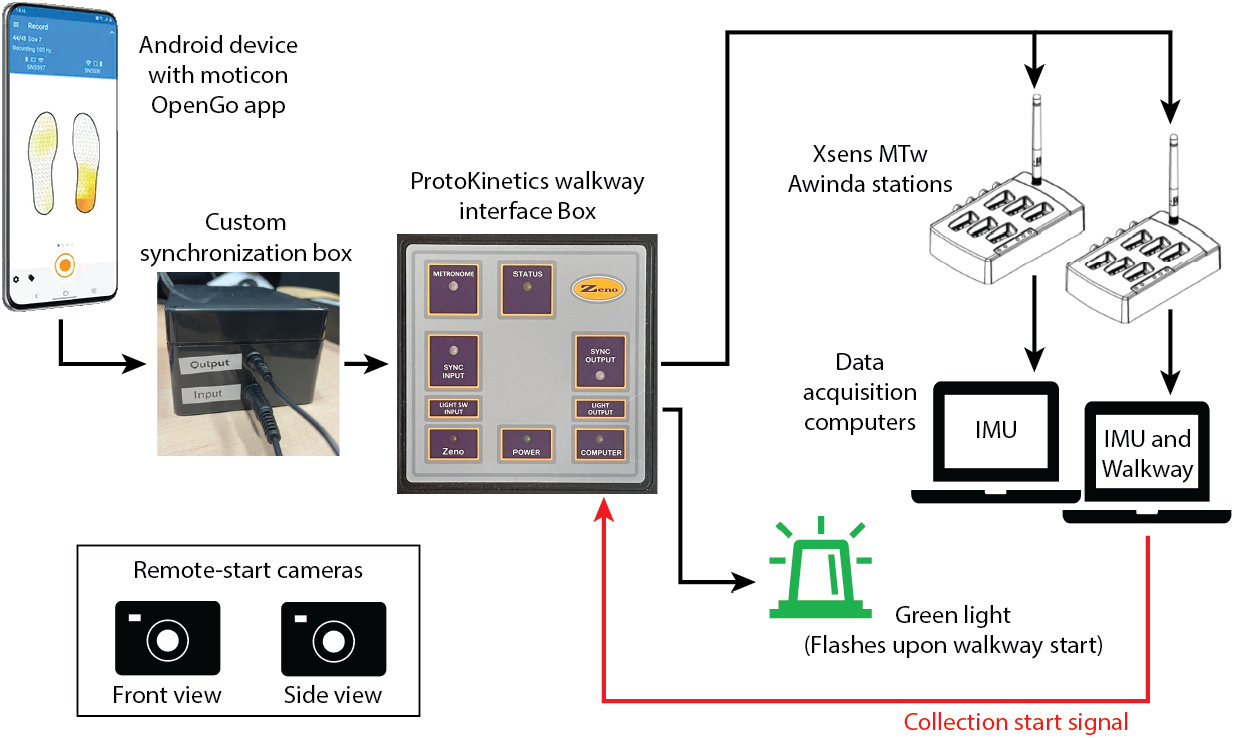
Synchronization approach across the walkway, IMU, sensor insole, and camera systems.

MATLAB scripts were written to read in the raw data from each sensor modality, align the data in time, and export to the MAT and CSV file formats. A brief description of the subroutines implemented to generate these aligned MAT and CSV files follows.

*Align walkway and IMU data →* Since the IMU system received a hardwired sync signal from the walkway system (primary clock) at the beginning of recording, the data from the IMU sensors and walkway were concatenated without further alignment. Occasionally, the IMU sensor data contained more or fewer time points than the walkway data. When shorter, padding of NaN was added to the end of the IMU data. When longer, the data that extended past the end of the length of the walkway data were discarded. Benchtop characterization confirmed the walkway and IMU data were aligned within 2 frames at 100 Hz.

*Align walkway and sensor insole data* → Custom electronics enabled a square-wave sync signal to be sent from the sensor insole system to the walkway system that denoted the start and stop of sensor insole recording. The end of the square wave (denoting sensor insole recording stop) was used as the alignment point as it was more consistent. The last frame of the sensor insole data was aligned with the last transition point of the sync square-wave in the walkway data. Padding of the beginning and end of sensor insole data with NaN was added to keep arrays the same size.

During pilot work, visual inspection of the aligned walkway and sensor insole data revealed a consistent time delay in the sensor insole signals relative to the walkway signals. We attributed this delay to the non-deterministic nature of the Android operating system running the Moticon Open Go App, which led to lag between stopping the sensor insole recording and the sync signal being sent to the walkway acquisition software. Benchtop experiments were conducted to characterize this delay across all study sites, sensor insole sizes, and data collection sessions. Results indicated site-specific, consistent delays that were independent of insole size and data collection session. These site-specific corrections were applied to the sensor insole data during initial import and alignment processing to address the constant bias component of the delay. However, cross-correlation analyses indicated that the delay between walkway and insole waveforms has both a constant component and small random components. Dataset users should proceed with caution when interpreting data that depends on precise alignment of the sensor insoles with the walkway and IMU systems. For relevant analyses, users may consider first aligning the insole total force waveforms to the walkway foot pressure waveforms using cross-correlation or other methods. More details on the system characterization can be found in the [Data Description – Sensor insole sync characterization](https://www.synapse.org/Synapse:syn52540892/wiki/623758) section of the Wiki.

*Align walkway and video annotation data* → The annotation file exported from MATLAB’s Video Labeler App was formatted such that the string variable for each annotation event at a given time point was listed under a ‘GeneralEvent’ or ‘ClinicalEvent’ column. The annotation data were resampled from 50 Hz to 100 Hz. Alignment between the walkway and video annotation data was achieved using the green light connected to the walkway system. A MATLAB script automatically detected the first video frame in which the green light turned on and used this to align the video annotation data with the start of the walkway data. All video data before the green light turned on were discarded.

The output of these processes is a set of aligned time-series data across each data collection source (Figure 4A), as well as the spatial information collected by the walkway (Figure 4B).

**Figure 4:**
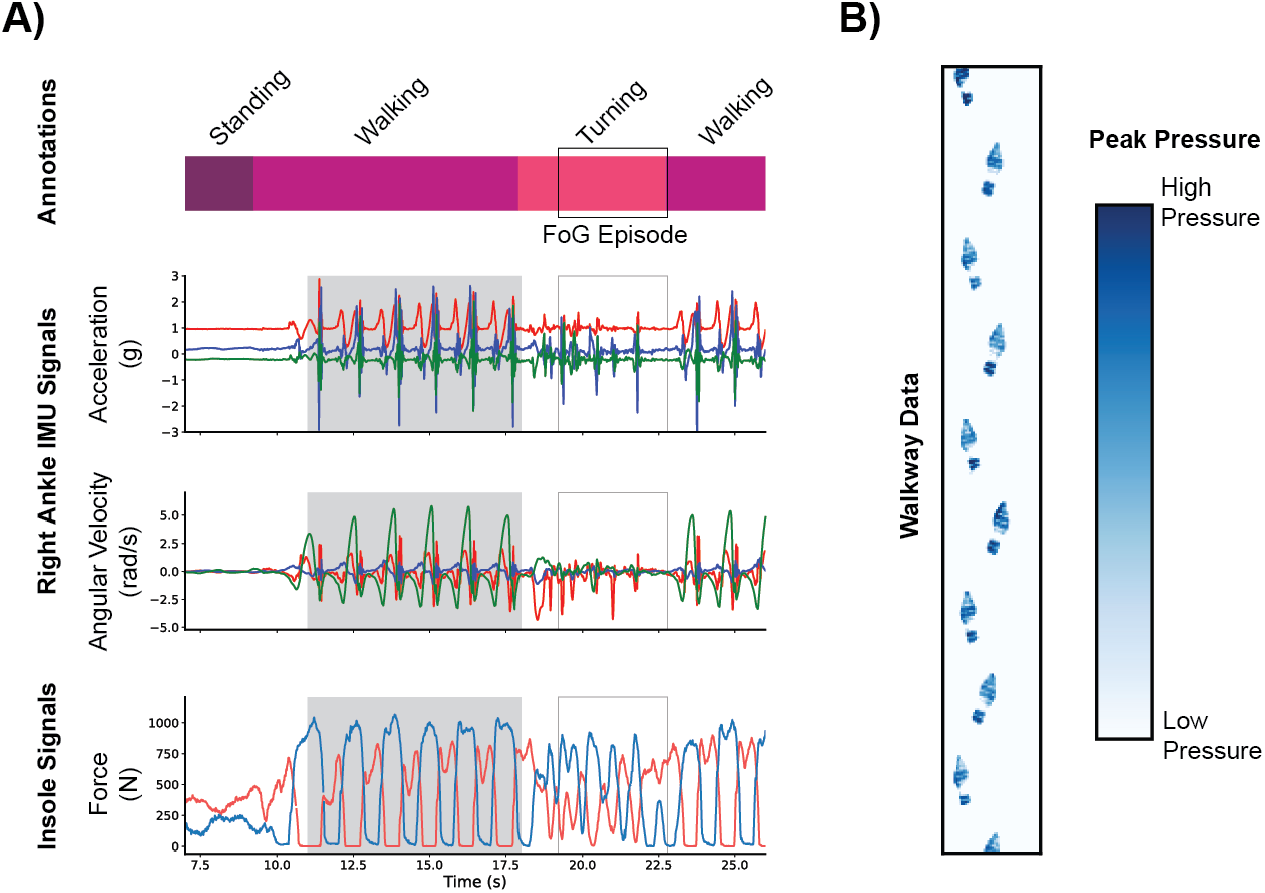
(A) Time series data from a participant with Parkinson’s disease, during which a freezing of gait (FoG) episode occurred. The data include annotation of activities captured via camera, 3-DOF accelerometer and gyroscope data from an IMU, and force data from a pair of sensorized insoles. The portion of the data with a gray background corresponds to the spatial information in part b. (B) Spatial data from a participant with Parkinson’s disease, during one pass across a sensorized walkway. Blue regions indicate areas of walkway activation, with darker colors corresponding to higher forces.

## Data Records

Detailed descriptions of the experimental set-up, data collection approach, processing, and data quality control for the WearGait-PD dataset are provided in the main Wiki page on the Synapse platform (SAGE Bionetworks) The [Data Access](https://www.synapse.org/Synapse:syn52540892/wiki/623752) tab within the Wiki provides further instructions on how to gain access to the data.

There are two main parts to the data: the clinical/demographic information and the sensor data. All clinical and demographic information is available for all PD patients and control participants and can be found in CSV spreadsheets. In these spreadsheets, each row constitutes one participant, with the first column listing the participant ID. All participant IDs are alphanumeric, with the letters denoting the study site at which the participant was recorded. All columns are labelled with the variable name. The clinical and demographic information contained within these spreadsheets was briefly described in the Methods – Clinical Information and Assessments section of this paper. When available, the response to each question of the MDS-UPDRS was included, allowing for maximum flexibility in use of the scores.

Formatting scripts were written to align all wearables and reference sensor data and save data as MAT files and comma-separated value (CSV) files. For a given participant, there are 8 CSV files (1 file/task) that contain the annotation events, walkway data, insole data, and IMU data. The CSV files follow the naming convention of <SubjectID>_<Task Name>.csv (see Methods – Tasks section of this paper for task descriptions). These data can be found in a folder labelled ‘CSV files’ in the Synapse repository. Of note, the use of the word ‘mat’ in the task names refers to participants performing a task entirely on the walkway mat.

The same data for each participant is also represented as one MATLAB MAT file that contains 8 structure variables for each task, with each task containing 8 fields (Figure 5).

**Figure 5:**
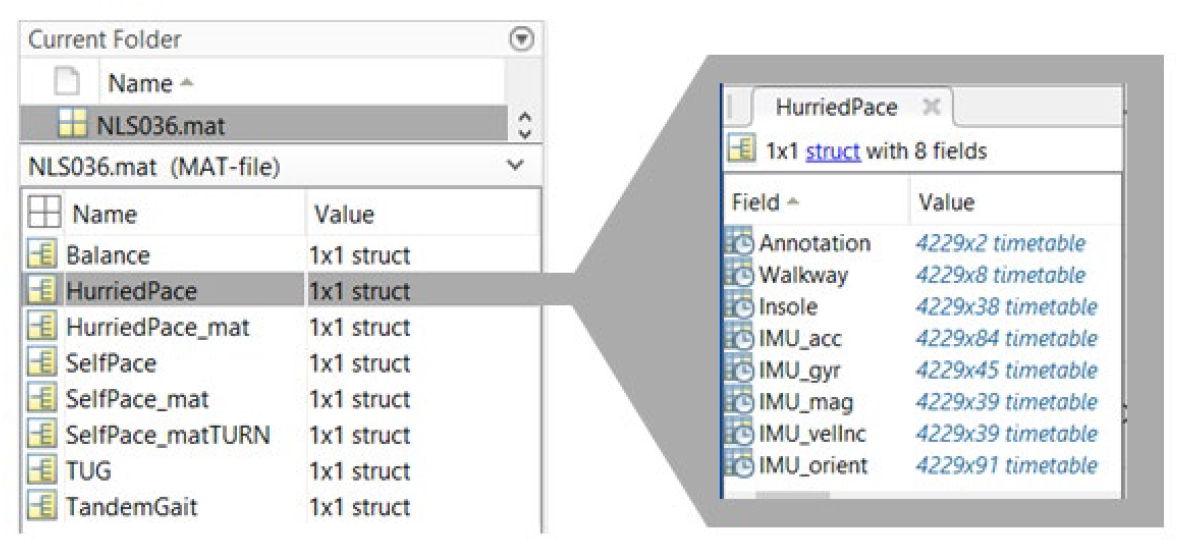
Sample MAT file structure for one participant.

A brief description of the fields within each task structure variable is below:

- Annotation: tx2 timetable containing video annotation information
- Walkway: tx8 timetable containing sensor and contact walkway data
- Insole: tx38 timetable containing insole data
- IMU_acc: tx84 timetable containing Acc and FreeAcc variables for each IMU and insole
- IMU_gyr: tx45 timetable containing gyroscope data for each IMU and insole
- IMU_mag: tx39 timetable containing magnetometer data for each IMU
- IMU_velInc: tx39 timetable containing Δv data for each IMU
- IMU_orient: tx91 timetable containing Δq and roll, pitch, yaw data for each IMU

For missing numeric data due to connectivity issues, sensor malfunctions, or lack of instrumentation for a given participant/task, NaN values were used. The data alignment process also used NaN values at the beginning and/or end to pad the time series, so NaN values in these locations are expected. Sensors not available during a given data collection were excluded from CSV output files but represented as NaN columns in the MAT files. For the GeneralEvent and ClinicalEvent variables containing frame-by-frame annotations, the string “unlabeled” was used to represent portions of a trial that did not match any annotation category (e.g., shuffling at the end of a trial instead of standing). In total, there were 346 variables associated with each task. A description of all the variables as well as reference coordinate systems for the IMUs, walkway, and sensor insoles is provided in the [Data Description](https://www.synapse.org/Synapse:syn52540892/wiki/623753) section of the Wiki.

## Technical Validation

Data quality is of the utmost importance in the curation of an open-access dataset. A quality assurance process was implemented to ensure consistent and accurate data collection. Specifically, a core experimental protocol was developed and reviewed by each researcher, including cross-site review of how the protocol was executed by various researchers. This protocol detailed the steps involved in system set-up, participant preparation, data collection, and data processing and export. All files were renamed to include the participant ID and task abbreviation immediately after data collection to minimize the risk of file loss due to incorrect folder allocation. For those modalities that required more manual processing (i.e., walkway data and video annotation data), a second researcher reviewed the primary processing completed by the first researcher to ensure proper footfall identification and conformance with the agreed upon annotation event definitions. An Excel spreadsheet accessible to all researchers was also used to track the progression of data processing, from initial collection to MAT and CSV file generation.

After MAT and CSV files were created, a data quality control process on those files was implemented through a series of automatic and manual data checks that involved visual inspection of data streams by a researcher to identify data cleanliness and validity issues.

Specifically, this process identified issues related to:

- Processing and inclusion of all tasks in the MAT and CSV files
- Integrity of individual IMU sensor data, including the identification of sensor values outside of expected ranges
- Unexpected missing columns of data
- Unexpected large gaps in data
- Expectations around the variable type for a given data variable (e.g., annotations contain no numeric data, IMU data does not contain any errant non-numeric data)
- Video annotation events and expectations surrounding the inclusion of specific events for specific tasks
- Alignment of all data, with a particular focus on walkway pressure and sensor insole force data

If an issue was identified, a second researcher was assigned to review and address the issue. Once an issue was addressed, the relevant files were put through the data quality control process again to confirm that all issues were resolved before the final MAT file and final set of CSVs files were generated and included in the dataset.

Of the 968 total trials that comprised the initial release of data, the majority of files passed all quality checks (651 trials) and another portion were flagged as potential issues but required no corrective action (58 trials) (Figure 6). Of the remaining trials with confirmed issues, 99 trials contained fixable issues such as the placement of files in the wrong location, missed annotation frames, or insole desynchronization. Each of these passed subsequent quality checks. The remaining 173 files had significant data loss (data dropout of 7 frames or greater, or loss of a sensor), either from connectivity or battery issues. While these data could not be recovered, the remaining data present in these trials were checked to confirm they were clean and valid. Overall, 82% of trials were considered complete, while the remaining 18% of trials passed all quality control checks with the exception that they were missing some data from either the insoles or IMUs.

**Figure 6:**
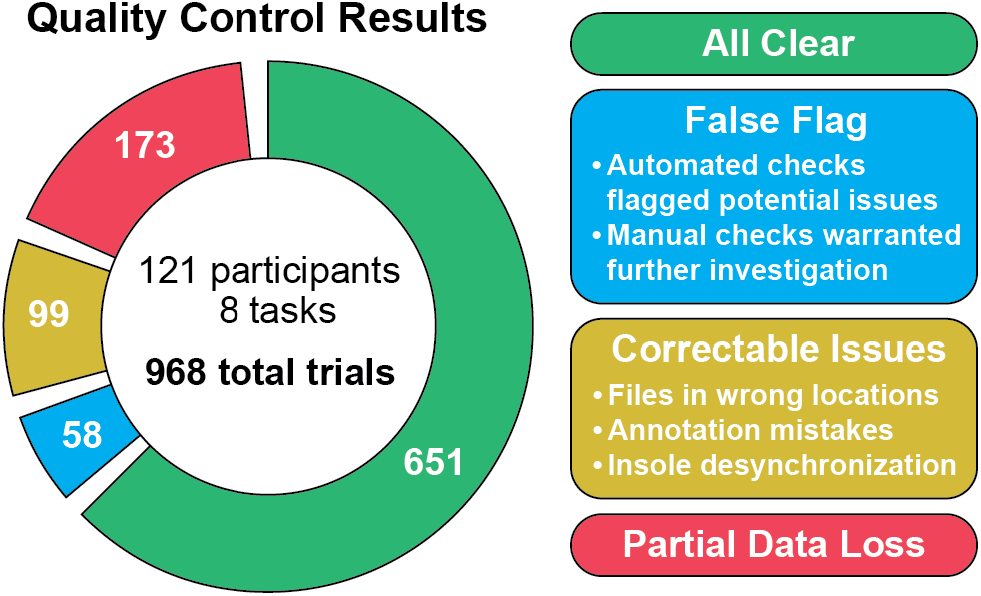
Quality control results for each trial, applied prior to inclusion of data to the WearGait-PD dataset. A trial is defined by a single task performed by a single participant. Category values add up to greater than 968 as the ‘False Flag’, ‘Correctable Issues’, and ‘Partial Data Loss’ categories were not mutually exclusive.

## Data Availability

All data produced are available online at https://www.synapse.org/Synapse:syn52540892/wiki/623751

## Usage Notes

To access the data on Synapse, users must first register with Synapse.org. Users register by providing a name and valid e-mail address and agreeing to the Synapse Terms and Conditions of Use and reviewing the Privacy Policy and Code of Conduct. As part of the registration process, users must agree to each of the terms of the Synapse Pledge, recapitulating the themes of the Synapse Terms and Conditions of Use, Privacy Policy, and Code of Conduct. Specifically, this pledge requires users to confirm they will (1) adhere to the community standards of inclusion and respect, (2) adhere to all conditions and data use limitations, (3) act ethically and responsibly, (4) use appropriate physical, technical, and administrative measures to keep data secure and protect participant’s privacy, (5) support open access best practices, (6) credit research participants and all data sources, (7) confirm the data will not be used for marketing and/or advertising, and (8) report suspected data breaches and/or misuse to the Synapse team.

## Code Availability

Using MATLAB 2023a, we developed code to implement the quality control process detailed in the Technical Validation section. This code is available in the ‘Code’ folder under ‘Files’ on the Synapse repository.

## Acknowledgements

The authors would like to thank all study participants for their enthusiastic participation. We’d also like to thank the FDA research fellows (Ms. Isabella Zuccaroli, Mr. Bryan Sabogal, Mr. Brian Nogh) who have assisted in data collection and processing, as well as former FDA staff members (Dr. Edward Nyman) who contributed to the development of the experimental protocol. The authors would also like to thank research coordinators from Johns Hopkins Bayview Medical Center (Ms. Jackie Langdon, Ms. Meredith Dobrosielski, and Ms. Crystal Szczesny) for their assistance with subject recruitment and consent, and Dr. Peter Abadir for his thoughtful insight into the experimental protocol. In this work, researchers at the Johns Hopkins University were partially funded by the Consolidated Anti-Aging Foundation.

## Author contributions

Conceptualization (KK, MC, AB), Methodology (All authors), Software (MC, MG, AA, KK), Validation (AA, MG, KK, MC), Formal Analysis (KK, AA, MG, DE), Investigation (KK, AA, DE, MG, NK, SW, MC, CM, BM, SZ, KM, AB, EM), Resources (KK, BM, AB), Data Curation (KK, AA, DE, MG, MC, NK, SW, SZ, AB, KM, EM), Writing – Original Draft (AA, DE, MG, KK), Writing – Review and Editing (All authors), Visualization (KK, AA, MG), Supervision (KK, BM, AB), Project Administration (KK, BM, AB, CM), Funding Acquisition (KK, BM, AA, LM, AB)

## Competing interests

The authors declare no competing interests.

### Disclaimer

The mention of commercial products, their sources, or their use in connection with material reported herein is not to be construed as either an actual or implied endorsement of such products by the Department of Health and Human Services.

